# Population Vaccine Effectiveness and its Implication for Control of the Spread of COVID-19 in the US

**DOI:** 10.1101/2021.04.30.21256228

**Authors:** Zixin Hu, Qiyang Ge, Li Luo, Tao Xu, Kai Zhang, Henry H Lu, Wei Li, Eric Boerwinkle, Li Jin, Momiao Xiong

**Affiliations:** State Key Laboratory of Genetic Engineering and Innovation Center of Genetics and Development, School of Life Sciences, Fudan University, Shanghai, China; Human Phenome Institute, Fudan University, Shanghai, China; The School of Mathematic Sciences, Fudan University, Shanghai, China; Department of Internal Medicine, University of New Mexico, USA; Department of Environmental Health Sciences, University at Albany, State University of New York, NY 12144, USA; Sanofi Pharmaceuticals. Bridgewater, NJ 08807; Department of Biostatistics and Data Science, School of Public Health, The University of Texas Health Science Center at Houston, Houston, TX 77030, USA

**Keywords:** Cov-19, vaccine, vaccine efficacy, vaccine distribution, forecasting

## Abstract

Realized vaccine efficacy in population is highly different from the individual vaccine efficacy measured in clinical trial. The realized vaccine efficacy in population is substantially affected by the vaccine age-stratified prioritization strategy, population age-structure, non-pharmaceutical intervention (NPI). We proposed a population vaccine efficacy which integrated individual vaccine efficacy, vaccine prioritization strategy and NPI to measure and monitor the control of the spread of COVID-19. We found that 11 states in the US had low population vaccine efficacy and 20 states had high population efficacy. We demonstrated that although the proportion of the population who received at least one dose of COVID-19 vaccine across 11 low population vaccine efficacy states, in general, was greater than that in 20 high population vaccine efficacy states, the 11 low population vaccine efficacy states experienced the recent COVID-19 surge, while the number of new cases in the 20 high population vaccine efficacy states exponentially decreased. We demonstrated that the proportions of adults in the population across 50 states were significantly associated with the forecasted ending date of the COVID-19. We show that it was recent low proportion of adults vaccinated in Michigan that caused its COVID-19 surge. Using population vaccination efficacy, we forecasted that the earliest COVID-19 ending states were Hawaii, Arizona, Arkansas, and California (in the end of June, 2021) and the last COVID-19 ending states were Colorado, New York and Michigan (in the Spring, 2022).

## Introduction

There is increasing hope for ending the spread of COVI-19 and returning normal through vaccination. On April 2, 2021, CDC reported that Pfizer-BioNtech and Moderna vaccines had 80% effectiveness against SARS-2 infection after first dose and 90% effectiveness after second dose (1). These vaccines were designed for the origin of SARS-CoV-2 virus. Unfortunately, mutations and natural selection generate new variants that increase virus replication, transmission, and escape of the immune system. There are three major rapidly circulating new variants: B.1.1.7 (that was initially reported in the United Kingdom on December 14, 2020), 501YV2 (B.1.351) (that was first announced in South Africa on December 18, 2020) and P.1(that was first described in Brazil on January 12, 2021), which reduce antibody neutralization and vaccine efficacy (2,3). These new variants raise concerns for increased transmission and escape from both vaccine and natural infection immunity.

It was reported that the Michigan coronavirus surged again. On April 6, 2021, the number of new cases in Michigan reached 6.041. However, on the same day, the total number of residents vaccinated was 3,054,258 and the coverage reached 37.7% (4). It was discovered that 40% of patients are infected with the B.1.1.7 variant.

Now the critical questions are whether the vaccine can end the COVID-19 epidemic, what causes the Michigan coronavirus to surge and how we can stop the spread of COVID-19? A key to correctly answer this question is vaccine distribution and dose prioritization plan design to increase population immunity levels against new variants. We identified the vaccine age distribution, which in turn caused the rapid transmission of B.1.1.7 variant in young population, was the major factor that led to the surge of Michigan coronavirus, using vaccine roll out real data and causal analysis. We developed a real data-based population vaccine efficacy index that took vaccine and non-pharmaceutical intervention (NPI) into account. We employ real-time COVID-19 transmission dynamic forecasting model and population vaccine efficacy index to design vaccine distribution curve and calculate the times when the COVD19 pandemic will end under the vaccine distribution plan for each state using the COVID-19 vaccination and new case data from January 12, 2021 to April 14, 2021, downloaded from https://ourworldindata.org/covid-vaccinations and https://coronavirus.jhu.edu/MAP.HTML.

## Results

Age-stratified vaccine distribution and prioritizing strategy were a major factor that caused the Michigan coronavirus surge again. To illustrate this, we presented Table 1 that showed that percentage of vaccinated adults 16-49 in Michigan was 29.94%, while percentage of vaccinated adults 16-49 in Texas was 39.97%. Vaccine age distributions between Michigan and Texas was significantly different (P-value < 10^−20^, Supplementary Materials, Materials and Methods). A given vaccine policy will have great impact on the continued spread of the new virus variants and COVID-19 as well (5). On January 6, 2021, no new variant was observed in Michigan, while the number of new variants in Texas was 1. However, on April 7, 2021, the number of new variants in Michigan increased to 1649, while the number of new variants in Texas increased to only 414. When COVD-19 surges shift to younger people due to lack of enough vaccination, the highly transmissible B.1.1.7 variant was rapidly circulating among young people, which in turn caused rises of COVID-19.

**Table 1.** Age distribution of vaccine receiving at least one dose in Texas and Michigan.

| Age | Texas |  | Michigan |  |
| --- | --- | --- | --- | --- |
|  | Total | Percentage | Total | Percentage |
| 16-49 | 3271084 | 0.3997 | 870493 | 0.2924 |
| 50-64 | 2362216 | 0.2886 | 902512 | 0.3031 |
| 65+ | 2551559 | 0.3117 | 1204373 | 0.4045 |

To investigate the relations between the new virus variants and the surge of COVID-19, we presented Figure S1 that plotted the new virus variant and new case of COVID-19 curves in Michigan and Texas. It was reported that on January 6, 2021, the number of new variant

(B.1.1.7) in Texas was 1, while the number of new variant in Michigan was zero (https://covariants.org/). However, on March 31, 2021, the numbers of new variants in Texas and Michigan were 414 and 1,237, respectively. Although the new variant B.1.1.7 appeared early in Texas than in Michigan, the increasing rate of new variants in Texas was smaller than that in Michigan. To further investigate whether the rises of the number of new cases of COVID-19 in Michigan was due to the new variant, we used the additive noise model to test the causation of the rapid increase of new cases by new variant (6). The P-values for new variant causation test in Texas and Michigan were 0.68 and 0.37, respectively. We did not observe significance of the causes of new cases by new variants. This showed that the new virus variants were not an original major risk for the rapid rise of new cases of COVID-19 in Michigan.

Both age-stratified vaccine distribution and NPI shaped the local transmission dynamics of COVD-19 before enough vaccination coverage was achieved. Vaccination efficacy that was determined in clinical trial should be adapted in allocation of a limited vaccine supply and real time forecasting transmission dynamics of COVID-19. The adapted vaccine efficacy should integrate vaccine efficacy, vaccine allocation strategy, and vaccine hesitancy and NPI (Supplementary Materials, Materials and Methods) (7). Figure 1 presented transmission dynamics and population vaccine efficacy curves in Texas and Michigan. We observed that when *α* coefficient was larger than 1 or, the population vaccine efficacy 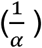 was less than 1, the number of new cases of COVID-19 began to increase in Michigan. More the a coefficient increased (or the population vaccine efficacy decreased), the more rapidly, the number of new cases of COVID-19 increased. However, in Texas, the vaccine efficacy stayed close to 1, the efficacy of vaccine was not affected or less affected by vaccine age distribution or intervention measure. The number of new cases of COVID-19 decreased exponentially when the proportion of vaccination in Texas increased. All *α* coefficients in 50 states were listed in Table 2 where a coefficients in each state were averaged over the observed period of time, starting with the time when the new case curve was in the recent valley. The population vaccine efficacy was significantly associated with the ending day of COVID-19 in 50 states with P-value < 1.25 × 10^−6^ (using Kendall’s rank correlation test) (8). The correlation coefficient between the population vaccine efficacy and the ending day was 0.4813. The ending day of COVID-19 in the state was defined as the day when the proportion of the number of new cases of COVID-19 was less than 3 × 10^−10^ or the number of new cases of COVID-19 was less than 10, while keeping the current vaccination distribution pace.

**Figure 1.**
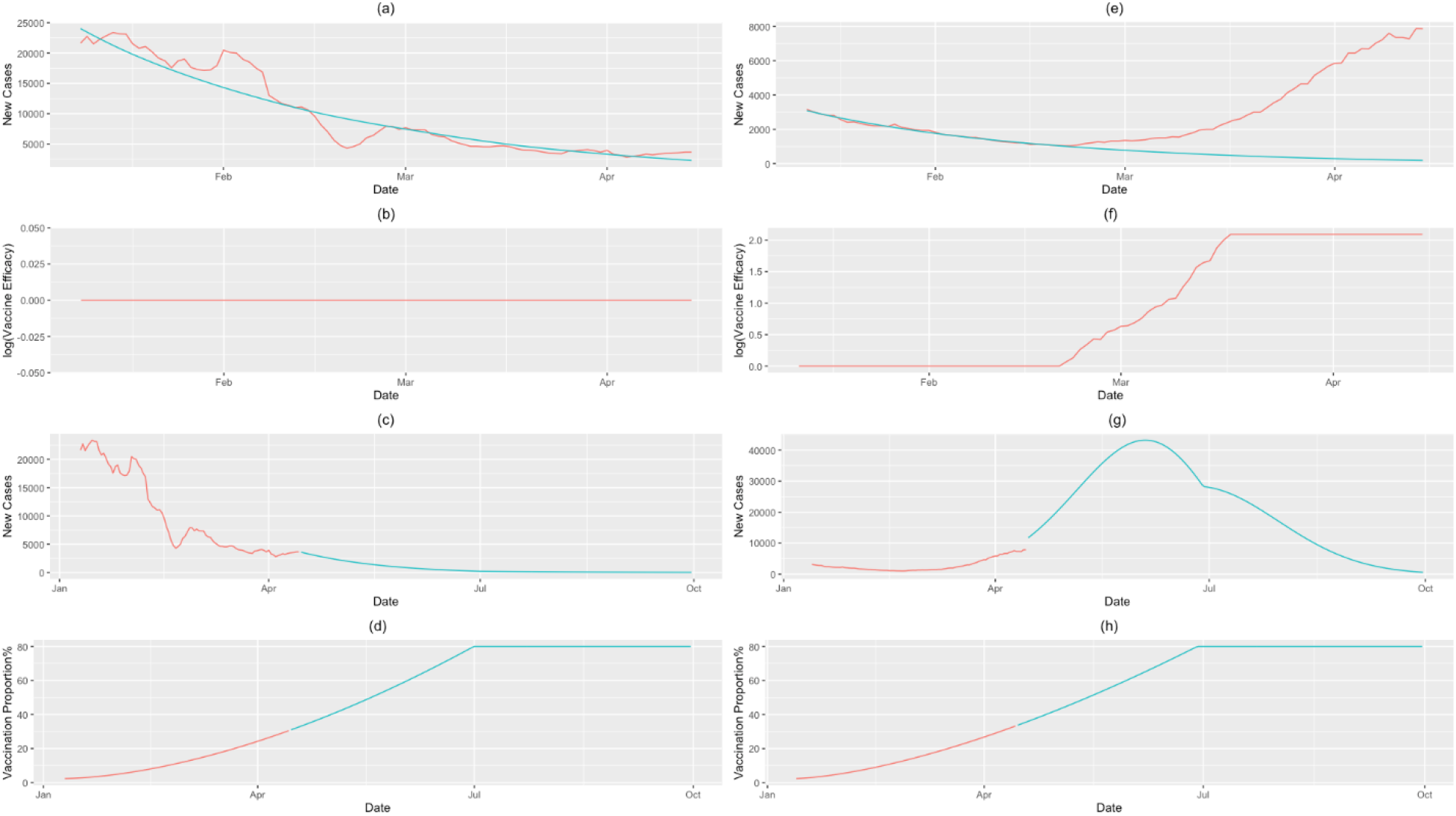
Transmission dynamics and population vaccine efficacy curves in Texas and Michigan. (a) New case curve in Texas where red curve represented the observed number of new cases of COVID-19 and green curve represented the fitted new case curve, (b) the curve of logarithm of the population vaccine efficacy and (c) the observed and forecasted new case curve in Texas where the red line was the observed and green line was the forecasted new case curves, vaccination proportion curve where red curve was observed and green curve was forecasted; New case curve in Michigan where red curve represented the observed number of new cases of COVID-19 and green curve represented the fitted new case curve, (f) the curve of logarithm of the population vaccine efficacy and (g) the observed and forecasted new case curve in Michigan where the red line was the observed and green line was the forecasted new case curves, (h) vaccination proportion curve where red curve was observed and green curve was forecasted.

**Table 2.** End day with current vaccine distribution rate and plan 1 to distribute vaccine in 51 states and regions.

| State | Current vaccine distribution rate | | | Vaccine Distribution Plan 1 | | | $\alpha$ | Gamma | 19-64 |
| --- | --- | --- | --- | --- | --- | --- | --- | --- | --- |
|  | End date | End cases | Prop of vaccine | End date | End cases | Prop of vaccine |  |  |  |
| Alabama | 8/1/2021 | 15 | 0.7149 | 8/1/2021 | 15 | 0.7149 | 1.00 | NA | 58.8 |
| Alaska | 8/14/2021 | 10 | 0.8000 | 12/6/2021 | 10 | 0.8000 | 1.87 | -9.9E-05 | 61.1 |
| Arizona | 6/25/2021 | 22 | 0.6876 | 6/25/2021 | 22 | 0.6876 | 1.81 | NA | 57.8 |
| Arkansas | 6/25/2021 | 9 | 0.6569 | 6/25/2021 | 9 | 0.6569 | 1.00 | NA | 58 |
| California | 6/27/2021 | 120 | 0.8000 | 6/27/2021 | 120 | 0.8000 | 1.00 | NA | 61.4 |
| Colorado | 3/28/2022 | 17 | 0.8000 | 3/28/2022 | 17 | 0.8000 | 13.26 | -4.4E-04 | 62 |
| Connecticut | 8/19/2021 | 11 | 0.8000 | 8/19/2021 | 11 | 0.8000 | 9.93 | NA | 60.6 |
| Delaware | 10/10/2021 | 3 | 0.8000 | 10/10/2021 | 3 | 0.8000 | 8.37 | NA | 58.5 |
| District of Columbia | 8/8/2021 | 2 | 0.8000 | 8/8/2021 | 2 | 0.8000 | 6.25 | NA | 67.8 |
| Florida | 10/23/2021 | 65 | 0.8000 | 10/23/2021 | 65 | 0.8000 | 5.08 | NA | 58.1 |
| Georgia | 8/19/2021 | 32 | 0.8000 | 8/19/2021 | 32 | 0.8000 | 1.00 | NA | 60.2 |
| Hawaii | 6/24/2021 | 4 | 0.7517 | 6/17/2021 | 4 | 0.8000 | 1.58 | -1.8E-04 | 57.5 |
| Idaho | 7/12/2021 | 5 | 0.7202 | 7/12/2021 | 5 | 0.7202 | 4.73 | NA | 57.1 |
| Illinois | 9/9/2021 | 38 | 0.8000 | 9/9/2021 | 38 | 0.8000 | 22.89 | -3.9E-04 | 60.3 |
| Indiana | 7/20/2021 | 20 | 0.6929 | 7/20/2021 | 20 | 0.6929 | 3.73 | NA | 59.3 |
| Iowa | 8/11/2021 | 10 | 0.8000 | 8/11/2021 | 10 | 0.8000 | 5.25 | NA | 58.2 |
| Kansas | 7/9/2021 | 9 | 0.8000 | 7/9/2021 | 9 | 0.8000 | 1.00 | NA | 57.9 |
| Kentucky | 9/2/2021 | 13 | 0.8000 | 9/2/2021 | 13 | 0.8000 | 1.00 | NA | 59.3 |
| Louisiana | 7/27/2021 | 14 | 0.7582 | 7/27/2021 | 14 | 0.7582 | 1.00 | NA | 59 |
| Maine | 10/30/2021 | 4 | 0.8000 | 10/30/2021 | 4 | 0.8000 | 20.62 | -3.9E-04 | 59.7 |
| Maryland | 7/20/2021 | 18 | 0.8000 | 7/20/2021 | 18 | 0.8000 | 11.31 | NA | 60.7 |
| Massachusetts | 9/2/2021 | 21 | 0.8000 | 9/2/2021 | 21 | 0.8000 | 9.29 | -3.3E-04 | 62 |
| Michigan | 1/14/2022 | 30 | 0.8000 | 9/26/2021 | 30 | 0.8000 | 8.10 | -4.1E-04 | 59.6 |
| Minnesota | 10/2/2021 | 17 | 0.8000 | 10/2/2021 | 17 | 0.8000 | 20.12 | -2.1E-04 | 59.5 |
| Mississippi | 7/22/2021 | 9 | 0.6377 | 7/22/2021 | 9 | 0.6377 | 1.00 | NA | 58.3 |
| Missouri | 7/8/2021 | 18 | 0.7487 | 7/8/2021 | 18 | 0.7487 | 2.71 | NA | 59.2 |
| Montana | 9/1/2021 | 3 | 0.8000 | 9/1/2021 | 3 | 0.8000 | 3.26 | NA | 57.8 |
| Nebraska | 8/8/2021 | 6 | 0.8000 | 8/8/2021 | 6 | 0.8000 | 9.39 | NA | 57.8 |
| Nevada | 7/31/2021 | 9 | 0.8000 | 7/31/2021 | 9 | 0.8000 | 3.37 | NA | 60.2 |
| New Hampshire | 11/9/2021 | 10 | 0.8000 | 11/9/2021 | 10 | 0.8000 | 13.43 | -6.7E-04 | 61.3 |
| New Jersey | 11/24/2021 | 27 | 0.8000 | 11/24/2021 | 27 | 0.8000 | 18.55 | -2.7E-04 | 60.5 |
| New Mexico | 8/22/2021 | 6 | 0.8000 | 8/22/2021 | 6 | 0.8000 | 1.85 | NA | 57.7 |
| New York | 3/2/2022 | 59 | 0.8000 | 3/2/2022 | 59 | 0.8000 | 10.01 | -3.8E-05 | 61.4 |
| North Carolina | 8/5/2021 | 32 | 0.8000 | 8/5/2021 | 32 | 0.8000 | 2.47 | NA | 59.6 |
| North Dakota | 12/12/2021 | 10 | 0.8000 | 10/10/2021 | 10 | 0.8000 | 22.14 | -1.6E-04 | 59.9 |
| Ohio | 7/30/2021 | 35 | 0.8000 | 7/30/2021 | 35 | 0.8000 | 4.30 | NA | 59.2 |
| Oklahoma | 7/27/2021 | 12 | 0.8000 | 7/27/2021 | 12 | 0.8000 | 1.00 | NA | 58.4 |
| Oregon | 8/4/2021 | 13 | 0.8000 | 8/4/2021 | 13 | 0.8000 | 9.19 | NA | 60.3 |
| Pennsylvania | 1/1/2022 | 38 | 0.8000 | 12/25/2021 | 39 | 0.8000 | 18.74 | -5.2E-04 | 59.4 |
| Rhode Island | 7/8/2021 | 10 | 0.8000 | 7/8/2021 | 10 | 0.8000 | 1.00 | NA | 61.5 |
| South Carolina | 9/21/2021 | 16 | 0.8000 | 9/21/2021 | 16 | 0.8000 | 1.00 | NA | 58.6 |
| South Dakota | 7/11/2021 | 3 | 0.8000 | 7/11/2021 | 3 | 0.8000 | **2.56** | -1.2E-04 | 56.9 |
| Tennessee | 8/1/2021 | 21 | 0.8000 | 8/1/2021 | 21 | 0.8000 | 2.47 | NA | 59.1 |
| Texas | 8/10/2021 | 87 | 0.8000 | 8/10/2021 | 87 | 0.8000 | 1.00 | NA | 59.9 |
| Utah | 8/15/2021 | 10 | 0.8000 | 8/15/2021 | 10 | 0.8000 | 1.00 | NA | 57.7 |
| Vermont | 11/28/2021 | 10 | 0.8000 | 11/28/2021 | 10 | 0.8000 | 8.45 | -3.1E-04 | 60.1 |
| Virginia | 8/3/2021 | 26 | 0.8000 | 8/3/2021 | 26 | 0.8000 | 3.31 | NA | 60.3 |
| Washington | 8/6/2021 | 23 | 0.8000 | 8/6/2021 | 23 | 0.8000 | 9.61 | NA | 61 |
| West Virginia | 8/11/2021 | 5 | 0.8000 | 8/11/2021 | 5 | 0.8000 | 3.09 | NA | 58.3 |
| Wisconsin | 8/22/2021 | 17 | 0.8000 | 8/22/2021 | 17 | 0.8000 | 6.29 | NA | 59.5 |
| Wyoming | 7/5/2021 | 10 | 0.6137 | 7/5/2021 | 10 | 0.6137 | 1.00 | NA | 57.7 |

Vaccination distribution rates and strategies determined when the spread of COVID-19 would be stopped. We considered four scenarios: (1) keep current vaccine distribution rate, (2) vaccine distribution plan1: on June 13, 2021, 80% of population will be vaccinated, (3) vaccine distribution plan 2: on May 30, 2021, 80% of population will be vaccinated, and (4) vaccine distribution plan 3: on June 27, 2021, 80% of population will be vaccinated. In Table 2 we summarized the ending date (that is defined as the day when the number of new cases of COVID-19 would be reduced to 3 × 10^−6^ of population), the number of new cases of COVID-19 and the proportion of vaccination at the ending date under the current vaccine distribution rate and vaccination distribution plan 1 in 50 states. Good news is that the spread of COVID-19 will be stopped in the US. The earliest ending day of COVID-19 will be 6/24, 2021 in Hawaii (6/24, 2021, the number of new cases will be reduced to 4 and 75% of population will be vaccinated), Arizona (6/25, 2021, the number of new cases will be reduced to 22 and 69% of population will be vaccinated), Arkansas (6/25, 2021, the number of new cases will be reduced to 9, 66% of population will be vaccinated) and California (6/27/2021, the number of new cases will be reduced to 120, 80% of population will be vaccinated). The last ending day will be 3/28, 2022 in Colorado (the number of new cases will be reduced to 17, 80% of population will be vaccinated), 3/2, 2022 in New York (the number of new cases will be reduced to 59, 80% of population will be vaccinated) and 1/14, 2022 in Michigan (the number of new cases will be reduced to 30, 80% of population will be vaccinated). By the end of august, 2021, majority of states (33 states) will almost stop the spread of COVID-19. By the end of this year, 46 states will contain the transmission of COVID-19. By the spring, 2022, all states and regions in the US will contain COVID-19, our life will go to normal.

When the population vaccine efficacy was reduced to 0.5, then the previous proportions of vaccination in some states were not large enough to prevent the rises of number of new cases of COVID-19. Half of the states in the US experienced no-decrease or even some surge of COVID-19 in the early of April, 2021. A non-negative parameter *γ* indicated the exponential increase of the number of new cases. There were 11 states with non-negative exponential growth parameter *γ*, including New Hampshire, Pennsylvania, Colorado, Michigan, Maine, Illinois, Massachusetts, New Jersey, Minnesota, North Dakota, and New York (Figures S2 and S5). The number of new cases of COVD-19 in these states increased recently. To further illustrate the importance of population vaccine efficacy for determining the transition dynamics of COVID-19 in the US, we presented Figure S3 showing the trajectory of observed and forecasted new cases of COVD-19 under current vaccination distribution rates and vaccine distribution plan 1 in 20 states with population vaccine efficacy larger than 0.3 (Table 2), Including South Carolina, Kentucky, Georgia, Utah, Texas, Alabama, Louisiana, Oklahoma, Mississippi, Kansas, Rhode Island, Wyoming, California and Arkansas, Arizona, New Mexico, Tennessee, North Carolina, Missouri and West Virginia. We also compared the proportions of adults (age 19-64) in these two groups.

The average proportion of adults in the group of states in which the number of new cases of COVID-19 recently decreased was 58.92% and the average proportion of adults in the group of states in which the number of new cases of COVID-19 recently increased was 60.52%. The Kendall correlation test showed that the proportion of adults in the state was significantly associated with the ending day of COVID-19 in the state (P-value < 0.00678).

The results for the vaccine distribution plans 2 and 3 were summarized in Table S1 and Figures S2, S3. Vaccine distribution plan 2 has some improvement on stopping transmission of COVID-19. The earliest ending states of COVID-19 will be Hawaii (5/27, 2021, the number of new cases will be reduced to 10, the proportion of vaccination will be 75.84%), District of Columbia (6/13, 2021, the number of new cases will be reduced to 10, the proportion of vaccination will be 76.83%), Arkansas (6/23. 2021, the number of new cases will be reduced to 10, the proportion of vaccination will be 64.56%), and Arizona (6/25, the number of new cases will be reduced to 22, the proportion of vaccination will be 68.76%). The last ending states under plan 2 will be New Jersey (4/7, 2022, the number of new cases will be 27, the proportion of vaccination will be 80%), Colorado (3/30, 2022, the number of new cases will be reduced to 17, the proportion of vaccination will be 80%) and New York (3/4, 2022, the number of new cases will be reduced to 58, the proportion of vaccination will be 80%).

Decreasing vaccine distribution rates will reduce the power of the vaccine on mitigating the spread of COVD-19 and delay its ending day. Since we only decreased small vaccination distribution rates, the results under vaccine distribution plan 3 only had small changes. In the list of early ending states, only the ending date in Hawaii was delayed from May, 27, 2021 to June 7, 2021 and the proportion of vaccination was changed from 75.84% to 66.75% of population.

There were no other changes in the list of early ending states. The list of last ending states added

Minnesota (4/27, the number of new cases was reduced to 17, the proportion of vaccination reached 80%) in it. The ending date in New Jersey was delayed from April 7, 2022 to April 14, 2022.

## Discussion

This study addressed several important issues in vaccination. We found that the age-structure of vaccine prioritization was an important factor that caused recent COVID-19 surge in Michigan. We compared the vaccine prioritization strategies for SARS-CoV-2 between Michigan and Texas and found that the proportion of vaccination in age group (16-49) in Michigan was much lower than that in Texas. We demonstrated by formal statistical test that difference in vaccination age-structure between Michigan and Texas was highly significant. It is less proportion of adults vaccinated in Michigan that caused its COVID-19 surge. The causal test also showed that the new virus variants were not a significant cause factor of the recent COVID-19 surge in Michigan. To further investigate the impact of age-structure on the spread of COVID-19, we compared the population age-structure across the 50 states in the US. We found that on the average, the proportion of adults in 11 low population vaccine efficacy states that experienced recent rises of COVID-19 were higher than that in 20 high population vaccine efficacy states with the recent decline of COVID-19. We showed that the proportions of the adults in the population was significantly associated with the ending date of COVID-19 in 50 states. The age structure may play an important role in the spread of COVID-19.

The efficacy of individual vaccine may not measure the real effect of vaccine in the population level. The vaccine prioritization strategies had a big impact on shaping local transmission dynamics. Also, the NPI plays a role in blocking the spread of COVID-19. The transmission dynamics of the COVID-19 is affected by the interplay between the NPI and vaccination. We proposed the population vaccine efficacy which integrate the NPI, vaccine distribution and vaccine efficacy to measure and monitor the control of the spread of COVID-19. We found that if the population vaccine efficacy in the state was close to one (the efficacy of individual vaccine to block the transmission of virus was not reduced in the population level), then the number of new cases decreased, while the average of population vaccine efficacies was less than 0.2, the number of new cases rapidly increased. The population vaccine efficacy was significantly associated with the ending date of COVID-19 in the State. The population vaccine efficacy determined the transmission dynamics of COVID-19 and hence was an important index for monitoring and controlling the spread of COVID-19.

COVID-19 vaccine was highly effective at stopping the spread of COVID-19. We forecasted that if we keep current vaccine distribution pace, the earliest ending date of COVID-19 would be in the end of June, 2021 (Hawaii, Arizona, Arkansas and California), and the last ending date would be in the end of March, 2022 (Colorado, New York and Michigan). In the Spring of next year, we may expect that the life in the US will return to normal.

## Data Availability

Data can be freely downloaded.

https://ourworldindata.org/covid-vaccinations

https://coronavirus.jhu.edu/MAP.HTML

## Author Contribution

Z. Hu, data analysis

Q. Ge, data analysis

L.Luo, Data analysis

T.Xu, data analysis

K. Zhang, data acquisition and interpretation

HH. Lu, concept and interpretation

W. Lin, supervision of analysis

E. Boerwinkle, biological interpretation

L. Jin, Concept and intepretation

M. Xiong, concept design, method development and write a manuscript

## ACKNOWLEDGEMENTS

Many thanks to the Texas Advanced Computing Center for computation support.

## Supplementary Figure

**Figure S1**. New virus variant and new cases of COVID-19 curves. The solid curves represent the new case curves and dotted curves represent the new variant curves. The curves in the red color represent Michigan and the curves in green color represent Texas.

**Figure S2**. Trajectory of observed and forecasted new cases of COVD-19 under current vaccination distribution rates and planned vaccination distribution rates in 11 states with non-negative exponential growth parameter y: New Hampshire, Pennsylvania, Colorado, Michigan, Maine, Illinois, Massachusetts, New Jersey, Minnesota, North Dakota, and New York. Curves in yellow, green, blue and pink colors represented the new case curves under current vaccine distribution rate, vaccine distribution plans 1,2 and 3, respectively.

**Figure S3**. Trajectory of observed and forecasted new cases of COVD-19 under current vaccine distribution rates and planned vaccine distribution rates in 20 high population vaccine efficacy states with population vaccine efficacy close to 1, Including South Carolina, Kentucky, Georgia, Utah, Texas, Alabama, Louisiana, Oklahoma, Mississippi, Kansas, Rhode Island, Wyoming, California, Arkansas, Arizona, New Mexico, Tennessee, North Carolina, Missouri and West Virginia. Curves in yellow, green, blue and pink colors represented the new case curves under current vaccine distribution rate, vaccine distribution plans 1,2 and 3, respectively.

**Figure S4**. Trajectory of observed and forecasted new cases of COVD-19 under current vaccination distribution rates and planned vaccination distribution rates in remaining 20 states in the US. Curves in yellow, green, blue and pink colors represented the new case curves under current vaccine distribution rate, vaccine distribution plans 1,2 and 3, respectively.

**Figure S5**. Population vaccine efficacy map where the states in green color, denoted their population vaccine efficacy larger than 0.3, the states in red color denoted their population vaccine efficacy smaller than 0.13 and with non-negative exponential growth parameter y, and the remaining states were in yellow color.

## References and Notes

1. Interim Estimates of Vaccine Effectiveness of BNT162b2 and mRNA-1273 COVID-19 Vaccines in Preventing SARS-CoV-2 Infection Among Health Care Personnel, First Responders, and Other Essential and Frontline Workers — Eight U.S. Locations, December 2020–March 2021. https://www.cdc.gov/mmwr/volumes/70/wr/mm7013e3.htm?s_cid=mm7013e3_w.

2. K. D. McCormick, J. L. Jacobs, J. W. Mellors. The emerging plasticity of SARS-CoV-2. Science, 371(6536):1306–1308 (2021).

3. S. S. Abdool Karim, T. de Oliveira. New SARS-CoV-2 Variants - clinical, public health, and vaccine Implications. N Engl J Med. doi: 10.1056/NEJMc2100362. (2021). Online ahead of print.

4. COVID-19 vaccine dashboard. https://www.michigan.gov/coronavirus/0,9753,7-406-98178_103214-547150--,00.html.

5. J. H. Buckner, G. Chowell, M. R. Springborn MR. Dynamic prioritization of COVID-19 vaccines when social distancing is limited for essential workers. Proc Natl Acad Sci U S 118(16):e2025786118 (2021).

6. R. Jiao, N. Lin, Z. Hu, D. A. Bennett, L. Jin, M. M. Xiong Mm. Bivariate Causal Discovery and its Applications to Gene Expression and Imaging Data Analysis. Front Genet. 9:347 (2018).

7. C. R. Wells, A. P. Galvani AP. The interplay between COVID-19 restrictions and vaccination. Lancet Infect Dis. S1473-3099(21)00074-8 (2021).

