## Supplementary Materials for "Population Vaccine Effectiveness and its Implication for Control of the Spread of COVID-19 in the US"

**Supplementary Material**

**Methods and Material**

**Test difference in vaccine distributions between two states**

Let $P_{j}^{A}, P_{j}^{B}, j=1,\ldots, m$ be the proportions of the vaccine distribution of the $j^{th}$ age group in the states $A$ and $B$, respectively, and $n_{A}, n_{B}$ be the total number of vaccinated individuals in the states $A$ and $B$, respectively .

Define the vaccine distribution vectors in the states $A$ and $B$:

$P^{A}=\left[ \begin{matrix} P_{1}^{A} \\ \vdots\\ P_{m-1}^{A} \end{matrix} \right]$ and $P^{B}=\left[ \begin{matrix} P_{1}^{B} \\ \vdots\\ P_{m-1}^{B} \end{matrix} \right]$.

Define the matrices:

$\Sigma_{A}=diag\left( P_{1}^{A}, \ldots, P_{m-1}^{A} \right)-P^{A}\left( P^{A} \right)^{T}$ and

$\Sigma_{B}=diag\left( P_{1}^{B}, \ldots, P_{m-1}^{B} \right)-P^{B}\left( P^{B} \right)^{T}$ .

The covariance matrix $\Lambda=cov(P^{A}-P^{B}, P^{A}-P^{B})$ is given by

$\Lambda=\frac{1}{n_{A}}\Sigma_{A}+\frac{1}{n_{B}}$ .

Define the test statistic:

$T=\left( P^{A}-P^{B} \right)^{T}\Lambda^{-1}\left( P^{A}-P^{B} \right)$ .

Under the null hypothesis of no vaccine distribution difference, $T$ is distributed as a central $\chi_{(m-1)}^{2}$ distribution.

If $m=2$, then $T$ is reduced to

$T=\frac{\left( P_{1}^{A}-P_{1}^{B} \right)^{2}}{\frac{P_{1}^{A}(1-P_{1}^{A})}{n_{A}}+\frac{P_{1}^{B}(1-P_{1}^{B})}{n_{B}}}$ .

Under the null hypothesis of no vaccine distribution difference, $T$ is distributed as a central $\chi_{(1)}^{2}$ distribution.

**Nonlinear additive noise models for bivariate causal discovery**

ANMs is used for identifying causal effect of a factor or an intervention measure on the number of new cases or an intervention measure (Peters et al. 2014; Jiao et al. 2018). Assume no confounding, no selection bias and no feedback. Let $Y$ be the average number of new cases of COVID-19 and $X$ be a potential causal factor. Consider a bivariate additive noise model
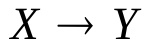
where
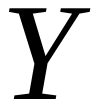
is a nonlinear function of
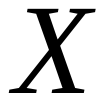
 and independent additive noise
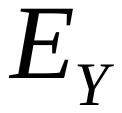
:

$Y=f_{Y}\left( X \right)+E_{Y}, X⫫E_{Y}$ ,	(S1)

where
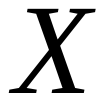
and
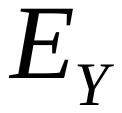
are independent. Then, the density $P_{X,Y}$is said to be induced by the additive noise model (ANM) from $X$ to $Y$ (Mooij et al. 2016). In some cases, we may have the following alternative direction ANMs:
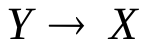
:

$X=f_{X}\left( Y \right)+E_{X}, Y⫫E_{X}$,	(S2)

where
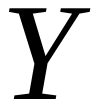
and
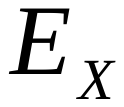
are independent. If the density $P_{X,Y}$is induced by the ANM
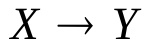
, but not by the ANM
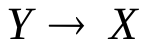
, then the ANM $X\to Y$ is identifiable.

Divide the $(n+m)$ sampled data points into a training data set by specifying $D_{1}=\left\{ Y_{n}, X_{n} \right\}, Y_{n}=\left[ y_{1, \ldots,}y_{n} \right]^{T} , X_{n}=\left[ x_{1},\ldots, x_{n} \right]^{T}$ for fitting the model and a test data set	$D_{2}=\left\{ \tilde{Y}_{m}, \tilde{X}_{m} \right\}, \tilde{Y}_{m}=\left[ \tilde{y}_{1},\ldots, \tilde{y}_{m} \right]^{T}, \tilde{X}_{m}=\left[ \tilde{x}_{1},\ldots,\tilde{x}_{m} \right]^{T}$	for testing the independence, where
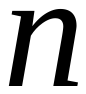
 is not necessarily equal to
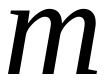
.

Procedures for using ANM to assess causal relationships between two variables are summarized below (Jiao et al. 2018).

**Step 1**. Regress $Y$ on $X$ using the training dataset $D_{1}$ and non-parametric regression methods:

$Y=\hat{f}_{Y}\left( X \right)+E_{Y}$.	(S3)

**Step 2**. Calculate residual $\hat{E}_{Y}=Y-\hat{f}_{Y}(X)$ using the test dataset $D_{2}$and test whether the residual $\hat{E}_{Y}$ is independent of causal $X$ to assess the ANM $\to Y$ .

**Step 3.** Repeat the procedure to assess the ANM $Y\to X$.

**Step 4**. If the ANM in one direction is accepted and the ANM in the other is rejected, then the former is inferred as the causal direction.

There are many non-parametric methods that can be used to regress
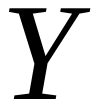
on
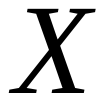
or regress
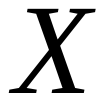
on
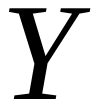
. For example, we can use neural networks (Heydari et al. 2019), smoothing spline regression methods (Wang 2011), B-spline (Wang 2017) and local polynomial regression (LOESS, see Cleveland , 1979). In this paper, smoothing spline regression method was used to fit the regression models.

The Hilbert-Schmidt norm of the cross-covariance operator or its approximation, the Hilbert-Schmidt independence criterion (HSIC) was used to measure the degree of dependence between the residuals and potential causal variable and test for their independence (Gretton et al. 2005; Mooij et al. 2016).

The Hilbert-Schmidt norm of the covariance operator can be used as criterion for assessing independence between two random variables and is called Hilbert-Schmidt independence criterion ${HSIC}^{2}(X,Y)$ .We know that (Wang et al. 2018)

${HSIC}^{2}(X,Y)=0$ if and only if $X$ and $Y$ are independent.

${HSIC}^{2}(X,Y)$ can be approximated by

${HSIC}^{2}\left( X,Y \right)=\frac{1}{n^{2}}\mathrm{tr}\left( KHLH \right)$ ,

where $n$ is a sample size, $K$ and $L=$ are $n\times n$ dimensional kernel matrices and $H=I-\frac{\boldsymbol{1}_{n}\boldsymbol{1}_{n}^{T}}{n}$. We used the Gaussian kernel: $k\left( x,y \right)=e^{-\frac{\left\| x-y \right\|^{2}}{2\sigma^{2}}}, \sigma>0$. To test independence between the potential cause $X$ and residual $E_{Y}$ , we calculated ${HSIC}^{2}(X,$ $E_{Y})$ as follows.

${HSIC}^{2}(X,$ $E_{Y})=\frac{1}{n^{2}}\mathrm{tr}(K_{X}HK_{E_{Y}})$,

$K_{E_{Y}}=\left[ \begin{matrix} k_{E_{Y}}(\varepsilon_{1}, \varepsilon_{1}) & \cdots& k_{E_{Y}}(\varepsilon_{1}, \varepsilon_{n}) \\ \vdots& \vdots& \vdots\\ k_{E_{Y}}(\varepsilon_{n}, \varepsilon_{1}) & \cdots& k_{E_{Y}}(\varepsilon_{n}, \varepsilon_{n}) \end{matrix} \right]$ and $K_{X}=\left[ \begin{matrix} k_{X}(x_{1}, x_{1}) & \cdots& k_{X}(x_{1},x_{n}) \\ \vdots& \vdots& \vdots\\ k_{X}(x_{n},x_{1}) & \cdots& k_{X}(x_{n},x_{n}) \end{matrix} \right]$,

$\varepsilon_{i}=\hat{E}_{Y(i)}=Y_{i}-\hat{f}_{Y}\left( X_{i} \right), i=1,\ldots,n,$

We calculate the dependence measures
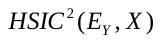
 and
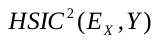
. Define a test statistic:

$T_{C}=|{HSIC}^{2}\left( E_{Y},X \right)-{HSIC}^{2}\left( E_{X},Y \right)|$.	(S4)

We do not have closed analytical forms for the asymptotic null distribution of the HSIC and hence it is difficult to calculate the P-values of the independence tests. To solve this problem, the permutation/bootstrap approach can be used to calculate the P-values of the causal test statistics. The null hypothesis is

$H_{0}:$ no causations $X\to Y$ and $Y\to X$ (Both $X$ and $E_{Y}$ are dependent, and $Y$ and $E_{X}$ are dependent).

Assume that the total number of permutations is
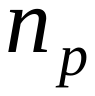
. For each permutation, we fix $X_{i,}i=1,\ldots, n$

and randomly permutate $Y_{i}, i=1, \ldots, n.$ Then, fit the ANMs and calculate the residuals $E_{X}^{i}, E_{Y}^{i}, i=1,\ldots, n$ and test statistic $T_{c}$. Repeat above procedures
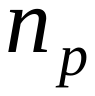
times. The P-values are defined as the proportions of the statistic $\tilde{T}_{C}$ (computed on the permuted data) greater than or equal to $\hat{T}_{C}$ (computed on the original test data $D_{2}$).

If the causation test is significant, then we infer causal direction:

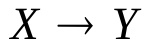
 if
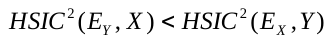
;	(S5)

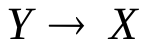
 if
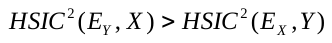
.	(S6)

If
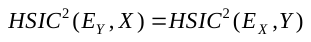
**,** then causal direction is undecided.

**Comprehensive Intervention Efficacy Integrating Vaccine Efficacy, Vaccine Allocation and non-Pharmaceutical Interventions**

Comprehensive intervention efficacy measures the effect of the vaccine and NPIs on mitigating the spread of COVID-19. Let $\omega(t)$ be a Comprehensive intervention efficacy at time $t$. Adapting the SIR model, we obtain

$\frac{dI(t)}{dt}=\beta\left( t \right)\left( 1-\omega\left( t \right)\nu\left( t \right) \right)\frac{S\left( t \right)}{N}I\left( t \right)-r\left( t \right)I(t)$ ,	(S7)

where $S\left( t \right).I(t)$ are the number of suspected and infected individuals, respectively, $\beta(t)$ and $r(t)$ are the exposure and recovery rates, respectively, $N$ is the total population, $\nu\left( t \right)$ is the proportion of the vaccinated individuals in the population. Let $R_{t}=\frac{\beta(t)}{r(t)}$ be the reproduction number at time $t$. Equation (S1) can be reduced to

$\frac{dI(t)}{dt}=\tau\left( t \right)I(t)$,	(S8)

where

$\tau\left( t \right)=r\left( t \right)[R_{t}\left( 1-\omega\left( t \right)\nu\left( t \right) \right)-1]$ .	(S9)

The solution to ordinary equation (S8) is given by

$I\left( t \right)=I(0)e^{\int_{0}^{t} \tau\left( x \right)dx}$ ,	(S10)

which implies

when $\omega\left( t \right)=\frac{1-\frac{1}{R_{t}}}{\nu(t)}$, the new case curve is in the peak or valley, when $\omega\left( t \right)<\frac{1-\frac{1}{R_{t}}}{\nu(t)}$, the number of new cases increases and when $\omega\left( t \right)>\frac{1-\frac{1}{R_{t}}}{\nu(t)}$, the number of new cases decreases. When the proportion of the vaccinated people $\nu(t$) is getting large, $\omega\left( t \right)>\frac{1-\frac{1}{R_{t}}}{\nu(t)}$ always holds. The number of new cases will finally decrease to zero.

In practice, the comprehensive intervention efficacy is calculated as follows. Let $y(t)$be the number of new cases of COVID-19 and $x(t)$ be the number of vaccinated individuals at time $t$. Assume that time $t_{*}$ is the closest to the time $t$, including time $t,$such that $y\left( t_{*} \right)=y(t)$. Let $x(t_{*})$ be the number of vaccinated individuals at time $t_{*}$. Define a comprehensive real time efficacy of the vaccine (or intervention) as

$\beta=\frac{x(t_{*})}{x(t)}$.

For the convenience of analysis, we define

$\alpha=\frac{1}{\beta}$ .

**The Decrease New Case Time Curve and Vaccination Time Curve**

Let $y(t)$ be the number of new cases of COVID-19, $V_{n}(t)$ be the number of the vaccinated people and $\nu(t$) be the proportion of the vaccinated people. Let

$y^{'}\left( t \right)=\frac{y(t)}{S-V_{n}(t)}$ .	(S11)

Define the exponential models for $y^{'}\left( t \right)$ and $\nu(t$):

$y^{'}\left( t \right)=ae^{-\tau_{y}t}$ ,	(S12)

$v\left( t \right)=\left\{ \begin{matrix} v_{0}e^{\tau_{v}t} & v(t)\leq v_{*} \\ v_{*} & v(t)\geq v_{*} \end{matrix} \right.$ .	(S13)

Use least square method, we can fit the exponential decrease or growth models for $y^{'}\left( t \right)$ and $\nu(t$).

Taking logarithm of both sides of equation (S5) yields

$\log y^{'}\left( t \right)=\log a-\tau_{y}t.$	(S14)

Let $z\left( t \right)=\log y'(t)$ and $b=\log a$. Equation (S8) is reduced to

$z\left( t \right)=b-t\tau_{y}$ .	(S15)

Use the least square method, we can solve the linear regression problem (S15). Then, the estimator of $y'(t)$ is given by

$y^{'}\left( t \right)=e^{z(t)}=ae^{-\tau_{y}t}$ .	(S16)

We can similarly use least square method to fit the vaccine-time curve.

The number of new cases $y(t)$ in the planned time period is estimated as

$y\left( t \right)=\left( 1-v\left( t \right) \right)Sy'(t)$ .	(S17)

**Increasing and then Decreasing New Case-Time Curve**

Consider second type of new case-time curve. The curve starts with increasing. After reaching the peak, it decreases. Let $t_{0}$ be the time when the curve begins. Let $y_{1}(t)$ be the number of new cases of COVID-19 in the second type of new cases-time curve. Define

$y_{1}^{'}\left( t \right)=\frac{y_{1}(t)}{S-V_{n}(t)}$ .	(S18)

Define the model for the second type of new case-time curve:

$y_{1}^{'}\left( t \right)=y_{1}^{'}\left( t_{0} \right)e^{[\tau_{c}-\gamma\left( t-t_{0} \right)](t-t_{0})}$ ,	(S19)

where $\tau_{c}$ and $\gamma$ are parameters in the model.

Let $z_{1}\left( t \right)=\log y_{1}^{'}\left( t \right)$. Taking logarithm on both sides of equation (S19), we obtain

$z_{1}\left( t \right)=\log y_{1}^{'}\left( t \right)+\tau_{c}\left( t-t_{0} \right)-\gamma\left( t-t_{0} \right)^{2}$ .	(S20)

Using least square method to fit the model (S20), we obtain the estimators $\hat{\tau}_{c}$ and $\hat{\gamma}$. The

the estimator of $y_{1}^{'}(t)$ is given by

$y_{1}^{'}\left( t \right)=e^{z_{1}(t)}=y_{1}^{'}\left( t_{0} \right)e^{[\hat{\tau}_{c}-\hat{\gamma}\left( t-t_{0} \right)](t-t_{0})}$ .	(S21)

The number of new cases $y_{1}(t)$ is estimated as

$y_{1}\left( t \right)=\left( 1-v\left( t \right) \right)Sy_{1}^{'}(t)$.	(S22)

**Supplementary Figure**

**Figure S1.** New virus variant and new cases of COVID-19 curves. The solid curves represent the new case curves and dotted curves represent the new variant curves. The curves in the red color represent Michigan and the curves in green color represent Texas.

**Figure S2.** Trajectory of observed and forecasted new cases of COVD-19 under current vaccination distribution rates and planned vaccination distribution rates in 11 states with non-negative exponential growth parameter $\gamma$: New Hampshire, Pennsylvania, Colorado, Michigan, Maine, Illinois, Massachusetts, New Jersey, Minnesota, North Dakota, and New York.

Curves in yellow, green, blue and pink colors represented the new case curves under current vaccine distribution rate, vaccine distribution plans 1,2 and 3, respectively.

**Figure S3.** Trajectory of observed and forecasted new cases of COVD-19 under current vaccine distribution rates and planned vaccine distribution rates in 20 high population vaccine efficacy states with population vaccine efficacy close to 1, Including South Carolina, Kentucky, Georgia, Utah, Texas, Alabama, Louisiana, Oklahoma, Mississippi, Kansas, Rhode Island, Wyoming, California, Arkansas, Arizona, New Mexico, Tennessee, North Carolina, Missouri and West Virginia. Curves in yellow, green, blue and pink colors represented the new case curves under current vaccine distribution rate, vaccine distribution plans 1,2 and 3, respectively.

**Figure S4.** Trajectory of observed and forecasted new cases of COVD-19 under current vaccination distribution rates and planned vaccination distribution rates in remaining 20 states in the US. Curves in yellow, green, blue and pink colors represented the new case curves under current vaccine distribution rate, vaccine distribution plans 1,2 and 3, respectively.

**Figure S5.** Population vaccine efficacy map where the states in green color, denoted their population vaccine efficacy larger than 0.3, the states in red color denoted their population vaccine efficacy smaller than 0.13 and with non-negative exponential growth parameter $\gamma$, and the remaining states were in yellow color.

**Table S1**

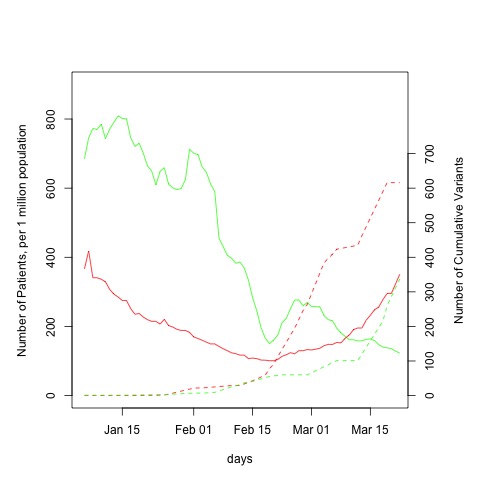

**Figure S1.** New virus variant and new cases of COVID-19 curves. The solid curves represent the new case curves and dotted curves represent the new variant curves. The curves in the red color represent Michigan and the curves in green color represent Texas.

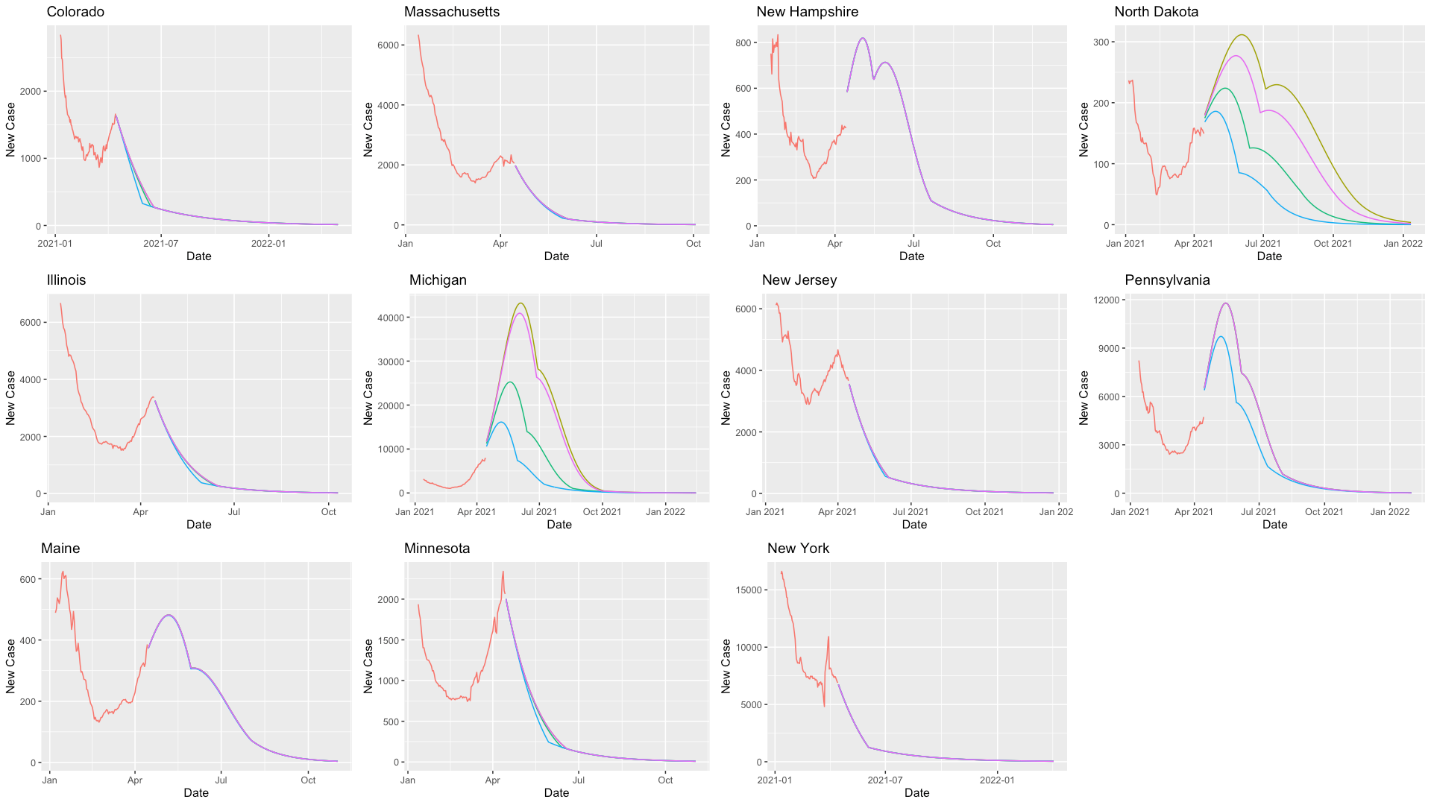

**Figure S2.** Trajectory of observed and forecasted new cases of COVD-19 under current vaccination distribution rates and planned vaccination distribution rates in 11 states with non-negative exponential growth parameter $\gamma$: New Hampshire, Pennsylvania, Colorado, Michigan, Maine, Illinois, Massachusetts, New Jersey, Minnesota, North Dakota, and New York.

Curves in yellow, green, blue and pink colors represented the new case curves under current vaccine distribution rate, vaccine distribution plans 1,2 and 3, respectively.

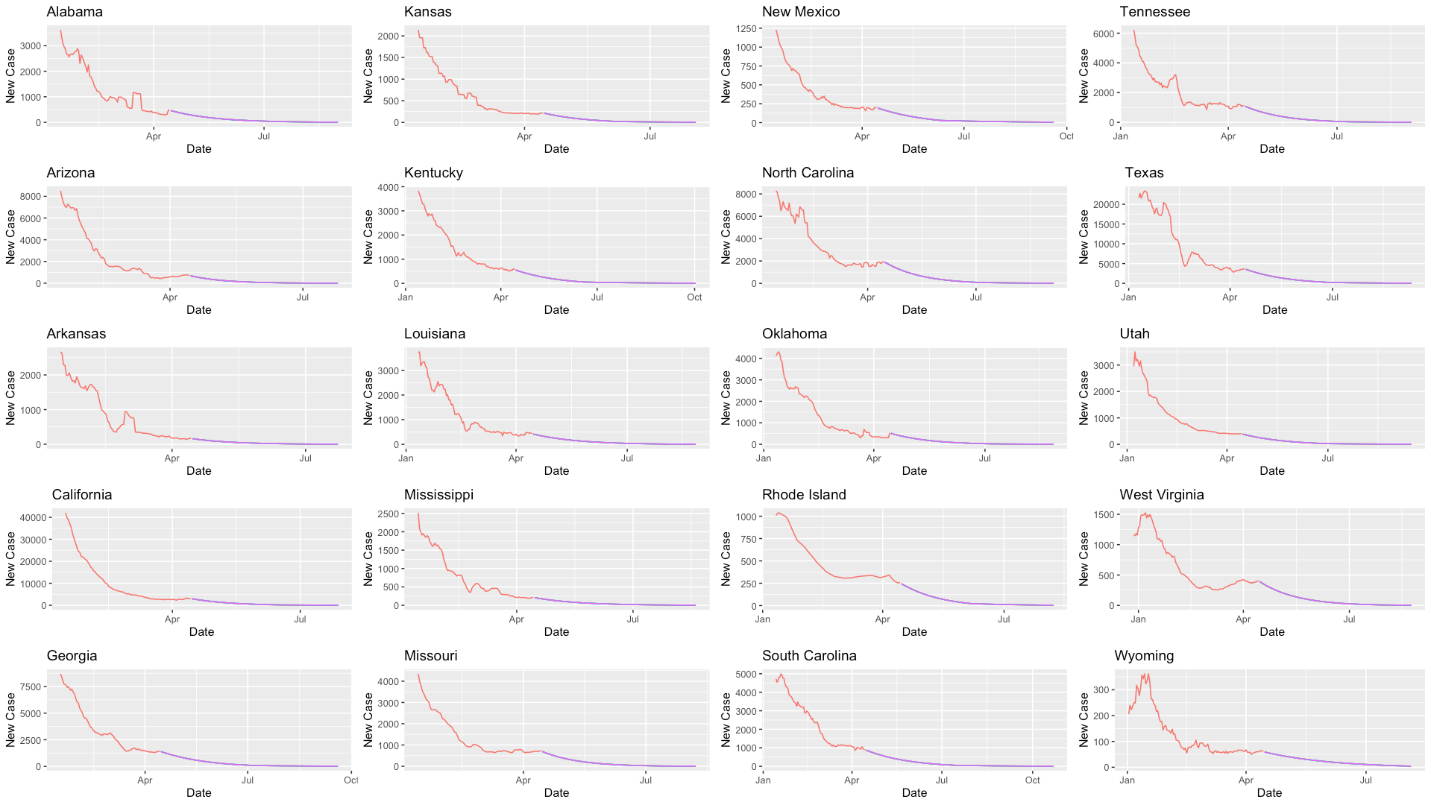

**Figure S3.** Trajectory of observed and forecasted new cases of COVD-19 under current vaccine distribution rates and planned vaccine distribution rates in 20 high population vaccine efficacy states with population vaccine efficacy close to 1, Including South Carolina, Kentucky, Georgia, Utah, Texas, Alabama, Louisiana, Oklahoma, Mississippi, Kansas, Rhode Island, Wyoming, California, Arkansas, Arizona, New Mexico, Tennessee, North Carolina, Missouri and West Virginia. Curves in yellow, green, blue and pink colors represented the new case curves under current vaccine distribution rate, vaccine distribution plans 1,2 and 3, respectively.

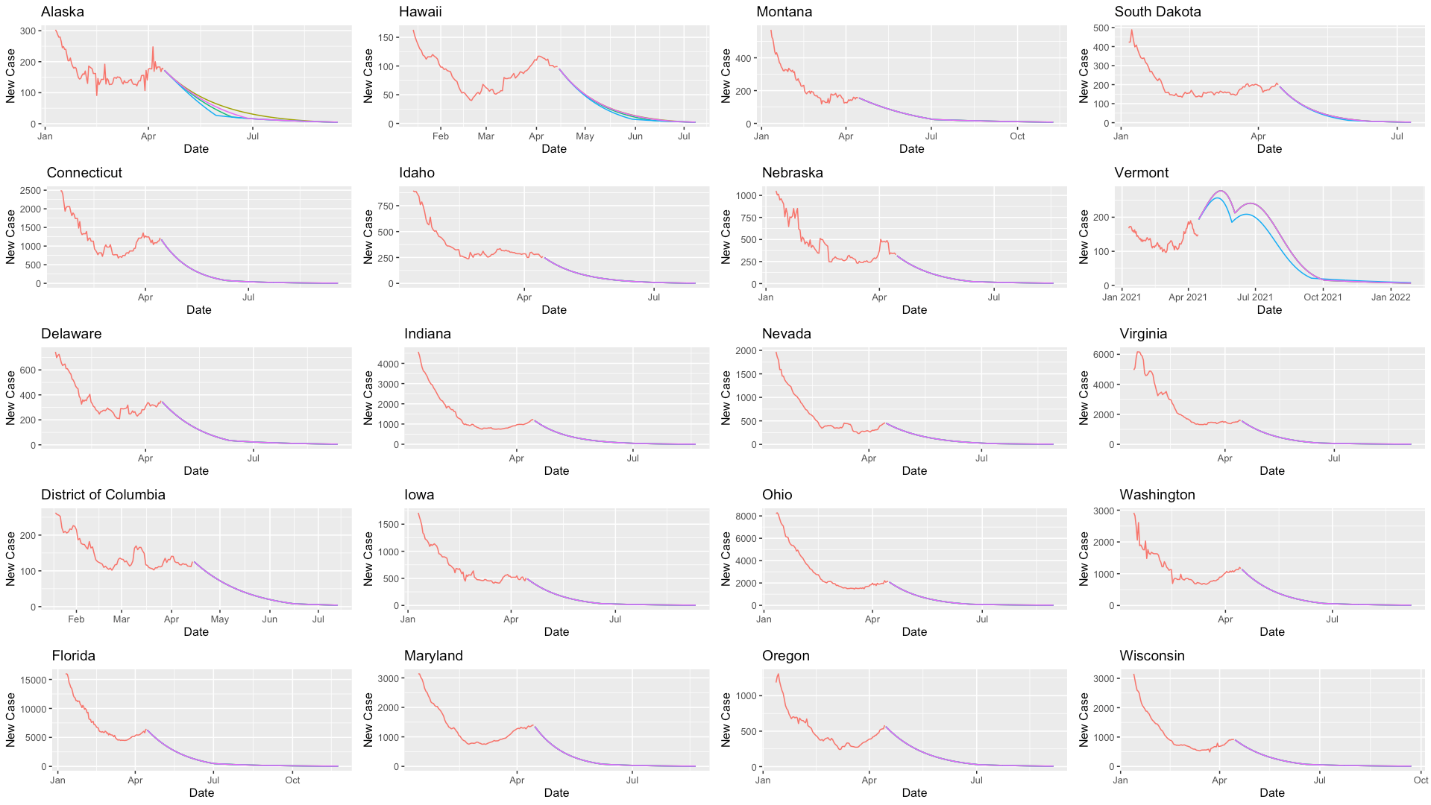

**Figure S4.** Trajectory of observed and forecasted new cases of COVD-19 under current vaccination distribution rates and planned vaccination distribution rates in remaining 20 states in the US. Curves in yellow, green, blue and pink colors represented the new case curves under current vaccine distribution rate, vaccine distribution plans 1,2 and 3, respectively.

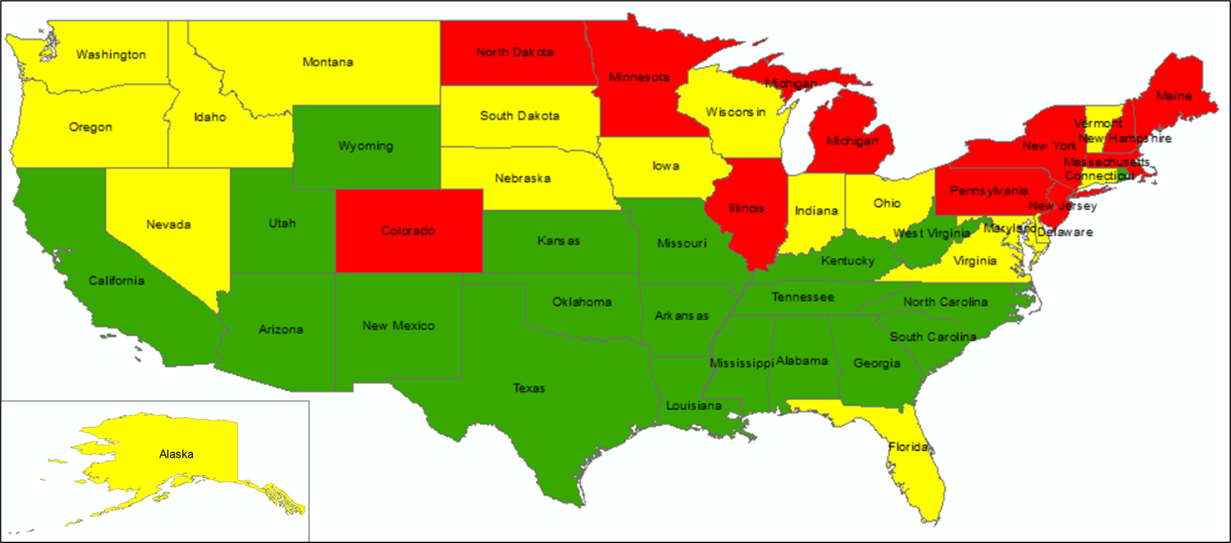

**Figure S5.** Population vaccine efficacy map where the states in green color, denoted their population vaccine efficacy larger than 0.3, the states in red color denoted their population vaccine efficacy smaller than 0.13 and with non-negative exponential growth parameter $\gamma$, and the remaining states were in yellow color.
